# Inferring the role of the microbiome on survival in patients treated with immune checkpoint inhibitors: causal modeling, timing, and classes of concomitant medications

**DOI:** 10.1101/19006429

**Authors:** Daniel Spakowicz, Rebecca Hoyd, Mitchell Muniak, Marium Husain, James S. Bassett, Lei Wang, Gabriel Tinoco, Sandip H. Patel, Jarred Burkart, Abdul Miah, Mingjia Li, Andrew Johns, Madison Grogan, David P. Carbone, Claire F. Verschraegen, Kari L. Kendra, Gregory A. Otterson, Lang Li, Carolyn J. Presley, Dwight H. Owen

**Author notes:** These authors contributed equally (Daniel Spakowicz, Rebecca Hoyd).

## Abstract

The microbiome has been shown to affect the response to Immune Checkpoint Inhibitors (ICIs) in a small number of cancers. Here, we sought to more broadly survey cancers to identify those in which the microbiome will play a role using retrospective analyses. We created a causal model for the relationship between medications, the microbiome and ICI response and used it to guide the abstraction of electronic health records of 690 patients who received ICI therapy for advanced cancer. Medications associated with changes to the microbiome including antibiotics, corticosteroids, proton pump inhibitors, histamine receptor blockers, non-steroid anti-inflammatories and statins were abstracted. We tested the effect of medication timing on overall survival (OS) and evaluated the robustness of medication effects in each cancer. Finally, we compared the size of the effect observed for antibiotics classes to taxa correlated with ICI response and a literature review of culture-based antibiotic susceptibilities. Of the medications assessed, only antibiotics and corticosteroids significantly associated with lower OS. The hazard ratios (HRs) for antibiotics and corticosteroids were highest near the start of ICI treatment but remained significant when given prior to ICI. Antibiotics and corticosteroids remained significantly associated with OS even when controlling for multiple factors such as Eastern Cooperative Oncology Group performance status, Charlson Comorbidity Index score, and stage. When grouping antibiotics by class, β-lactams showed the strongest association with OS across all tested cancers. The timing and strength of these effects after controlling for confounding factors are consistent with role for the microbiome in response to ICIs.

## BACKGROUND

Treatment with Immune Checkpoint Inhibitors (ICIs) has improved patient outcomes across a wide variety of cancers. Not all patients respond to these drugs and there is a need to identify biomarkers of outcomes. Three recent studies have shown that microbes are associated with response and overall survival (OS) in renal cell carcinoma (RCC), non-small cell lung cancer (NSCLC) and melanoma (1–3). The microbiome may be a key player in response to ICI therapy and a potential biomarker of treatment response.

The microbiome is known to interact with the immune system, but how it affects response to ICIs is not understood. The effectiveness of ICI treatment relies on active T-cell infiltration of a tumor; microbes have been associated with increased Tumor Infiltrating Lymphocytes in an IL12-depended manner (4). However, other immune cells dampen response to ICIs such as myeloid-derived suppressor cells and FOXP3 & CD4+CD25+ T-regulatory cells. Another, systemic form of immune repression is characterized by the production of prostaglandins (5–7).

Several medications commonly used during routine oncologic care and ICI treatment can influence inflammation pathways and/or the microbiome. Corticosteroids (CS) affect both of the aforementioned T-cell subtypes and the prostaglandin-related inflammatory pathways (8). Additionally, antibiotics (ABx) have a direct effect on the microbiome by killing or halting the growth of bacteria. Proton pump inhibitors (PPIs), histamine 2 blockers (H2Bs), non-steroid anti-inflammatory drugs (NSAIDs), and CS have also been associated with changes in the microbiome but, in contrast to antibiotics, this mechanism is indirect (9). PPIs, by inhibiting gastric acid secretion, alter the pH of the gut and change the number and types of bacteria that pass through the stomach (10). Notably, if the taxa enriched by the PPI-induced pH change are also important for response to ICIs, then PPI treatment during ICI may influence clinical outcomes. The effect of other medications on clinical response may be challenging to interpret given that the effects may influence both the microbiome and ICI response.

In order to disentangle these complex interactions, we created a model of the relationship between patient characteristics, medications that affect the microbiome, inflammation, and survival. Second, we performed a retrospective analysis of patients who received ICI therapy for advanced cancer between 2011 and 2017 including medications with known effects on either the microbiome or its pathway toward affecting ICI response. Third, we estimated the association for each medication with OS. Fourth, we analyzed the effects of medications longitudinally, in order to decouple confounding variables at different time points. Fifth, we controlled for variables that broadly describe differences in baseline statuses (e.g. Eastern Cooperative Oncology Group performance status (PS)) of individuals who received concomitant medications and those who did not. Sixth, we compared the associations across several cancers, for which the medications are prescribed in subtly different ways that can be leveraged to gain further insight into the causal effects. Finally, we related these results to the microbes shown to be enriched or depleted in individuals who respond to ICIs. The combination of these strategies gives layers of support to defining the role of the microbiome in the context ICI treatment of cancer.

## METHODS

### Causal Model

We performed a literature review of the relationship between the microbiome and response to ICIs and medications that affect the microbiome (**Figure 1**, references in **Figure S1**). From these references, a causal model was then constructed such that the nodes correspond to observable endogenous variables (*Vi*), as a subset of a set of *U* exogenous and unobserved variables that affect the relationship between the microbiome and OS in patients treated with ICIs. Directed edges denote a relationship between variables when the following conditions are met: (1) there is a reported relationship between variables in which both variables were either observed or defined by intervention, and (2) the relationship cannot be explained through using an existing path. For example, Gopalakrishnan et al reported a correlation between the microbiome and ICI response (1). This relationship exists in the graph as mediated by the nodes Microbiome → T-cell Mediated Inflammation → ICI Response, therefore no edge is drawn directly from Microbiome → ICI Response. The resulting directed acyclic graph was constructed using the igraph and dagitty packages in R (11,12).

**Figure 1.**
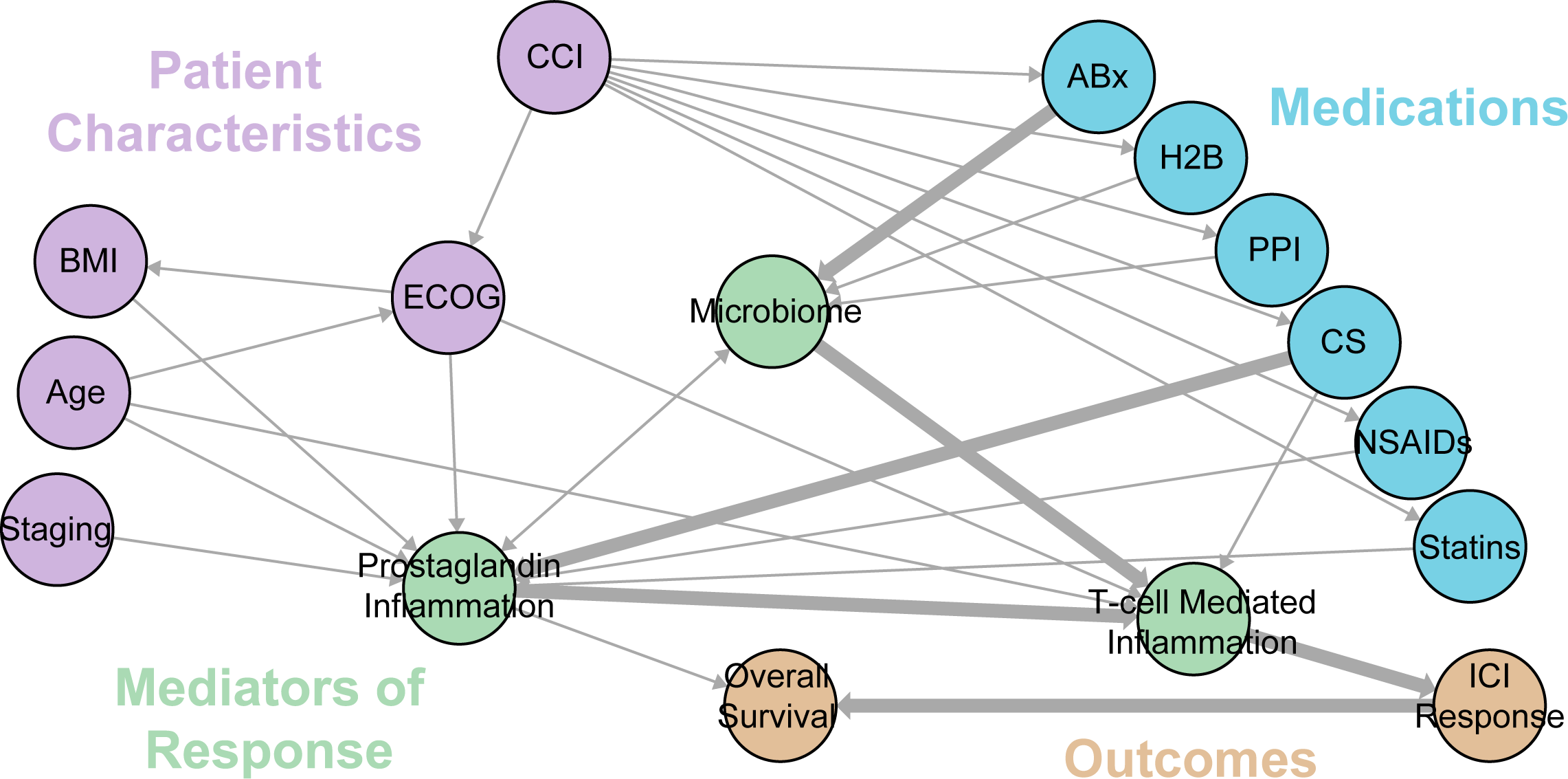
Causal model for the effect of concomitant medications on Immunotherapy Response and Overall Survival. Numbers along edges refer to references supporting the connection. Hypothesized dominant pathways are shown in heavily-weighted edges. (1,2,8–10,27–29,31,32,36–52)

### Retrospective data collection

We identified patients with advanced cancer treated between 2011 and 2017 at the Ohio State University Comprehensive Cancer Center/Arthur G. James Cancer Hospital (OSUCCC-James) who received at least one dose of ICIs as part of an IRB approved retrospective study (OSU-2016C0070, OSU-2017C0063). Patient data were collected and housed in REDCap (13). Medication timing, dose and names were collected from the electronic medical record information warehouse and validated by manual chart review. Additional diagnoses prior to ICI start were manually recorded from the Problem List, Medical History, and Encounter Diagnoses in the electronic medical record and compiled using the Charlson Comorbidity Index (CCI) (14), which includes record of myocardial infarction, congestive heart failure, peripheral vascular disease, cerebrovascular disease, dementia, chronic pulmonary disease, connective tissue disease, ulcer disease, mild liver disease, diabetes, hemiplegia, moderate or severe renal disease, moderate or severe liver disease (e.g., cirrhosis with ascites), or HIV AIDS.

### Medication history curation

ABx and CS data were retrieved from the information warehouse within 180 days of ICI start. All medications matching a comprehensive list of steroid generic and brand names were collected with dates and routes of administration. Medications were filtered to those confirmed to be administered and the results checked against a manually-curated subset of the records.

### Survival Analysis

Overall survival (OS) was reported in days from the initiation of ICI to the date of death or last follow-up. All univariate and multivariate analyses were conducted using the survminer package in R (15,16). Univariate analyses used Kaplan-Meier survival curves with log-rank tests. Multivariate analyses used Cox-Proportional Hazards models, defining the hazard function for each patient *k* as:

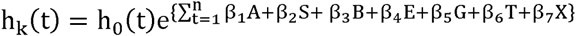

Where h(t) is the hazard function at time t = 1 to *n*, ***A*** is a binary indicator of antibiotic use (+/- 28 days from start of ICIs), ***S*** is a binary indicator of corticosteroid use (+/- 28 days from start of ICIs), ***B*** is BMI, ***E*** is the Eastern Cooperative Oncology Group performance status score (1-5), ***G*** is age, ***T*** is stage and ***X*** is sex. We constructed the models using the survival package and evaluated model fits using a likelihood ratio tests in R (17–19).

### Timing analysis

A 30-day sliding window was used to evaluate the effect of medication timing on the association with OS. Patients prescribed medications within the window were compared to a cohort of individuals who were not prescribed those medications within 180 days before or after the start of ICI treatment. Kaplan-Meier survival curves were used to estimate a hazard ratio (HR) of association with each treatment window, incremented by single-days, e.g. prescribed 180-150 days before ICI start vs no prescribed medications, and then prescribed 179-149 days before ICI start vs no prescribed medications. HRs and confidence intervals were calculated in the survival package and plotted with ggplot2 in R (18–20).

### Antibiotics and Corticosteroids classes

ABx and CS were collapsed into categories by DrugBank v5.0 accession numbers (21). HRs were estimated for medication class and cancer combinations if the total sample set included at least 20 individuals. Cox Proportional Hazards models for the effects of ABx and CS class were used to allow for simultaneous estimation of the effects of more than one class, when applicable. Plots showing prescriptions of more than one class were created with the UpSetR package in R (22).

### Regularized Cox Regression

Regularized Cox survival models for each cancer were implemented in the glmnet and coxnet packages in R (23,24). We optimized the regularization parameter by coordinate descent via 10-fold cross-validation and then tested the robustness of the parameter selection and resulting covariates with 1000 bootstrap replicates of different random samples of the dataset (23,24).

### Reproducibility

Scripts to reproduce all figures and analyses can be found at https://github.com/spakowiczlab/co-med-io.

## RESULTS

### Causal Model

The relationships between clinical variables, medications, the microbiome, ICI response and OS are strongly interconnected. Our literature review to predict their relationships (**Figure 1**) led to several hypotheses testable within retrospective data. First, medications that affect ICI response via the microbiome will proceed through T-cell mediated inflammation (i.e. ABx → Microbiome → T-cell mediated inflammation → ICI Response → OS). Second, the use of these medications is driven by comorbidities, which must be controlled for. Here we attempt this using the Charlson Comorbidity Index (CCI) to capture and simplify several disease states (25). Non-ICI-response effects on OS proceed through Prostaglandin Inflammation. For example, the path CCI → ABx→ Microbiome → Prostaglandin Inflammation → OS may include sepsis, through which inflammatory processes may lead to low blood pressure or multi-system organ failure and therefore OS. Third, CS and ABx may have additive effects on ICIs through a collider effect on T-cell mediated inflammation (i.e. ABx → Microbiome → T-cell mediated inflammation ← CS). Finally, the clinical variables of stage, BMI, and age, and medications such as CS confound the relationship between the microbiome and ICI response, mediated by Prostaglandin-based inflammation (which itself is a collider), and therefore must be controlled for in order to infer the role of the microbiome on OS (**Figure 1**).

### Patient Characteristics

Retrospective analysis of electronic medical records from 2011 to 2017 at the OSUCCC-James identified 690 patients treated with ICIs (**Table 1**). Most (76.6%) had a PS of 0 or 1 and 0-1 co-morbidities (CCI 0-1, 66.7%). The most common diagnoses were melanoma (28.5%) and non-small cell lung cancer (NSCLC) (23.4%). Cancers represented by fewer than 20 patients were categorized as “Other” (23.4%). The majority of patients (90%) had metastatic disease. ICI treatments included nivolumab in 52.8% of patients, ipilimumab in 18.0% and pembrolizumab in 15.1%.

**Table 1.**
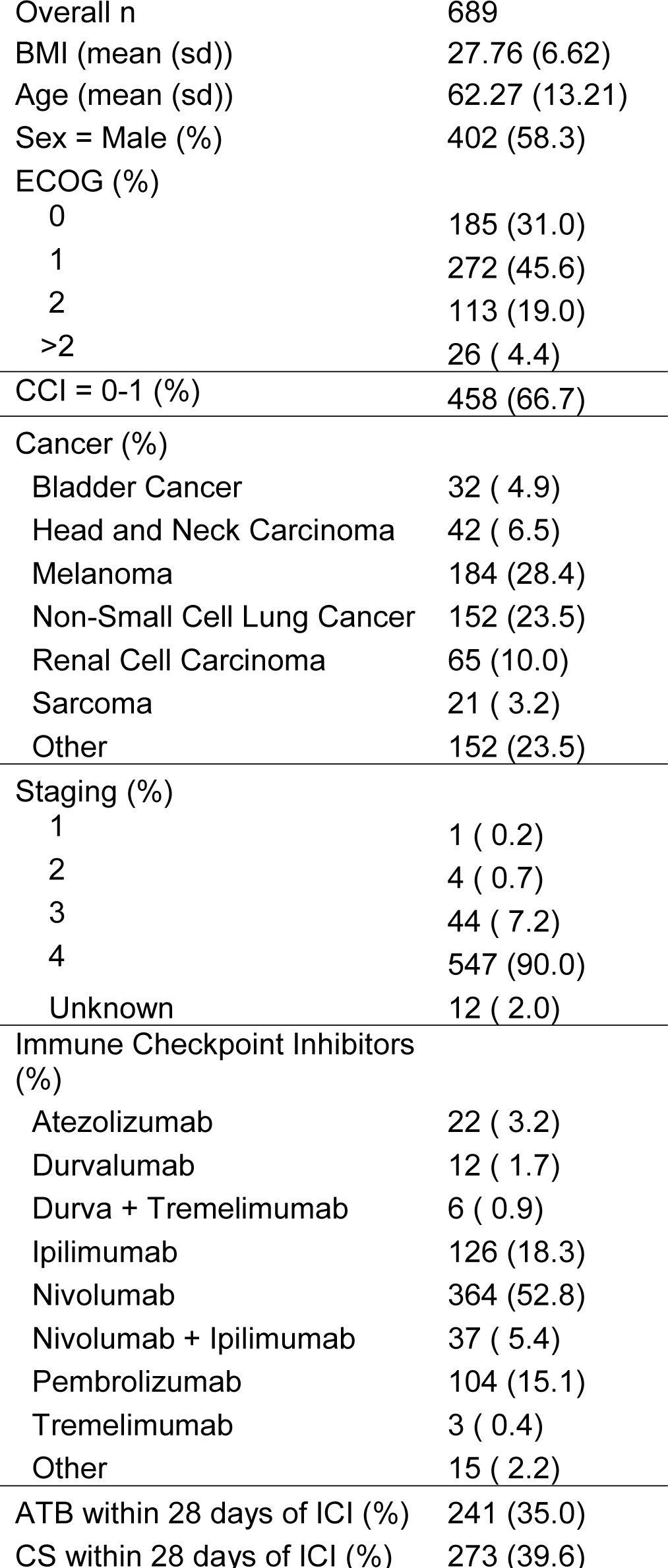
Cohort characteristics

### Microbiome and inflammation-related concomitant medication use

Among the medications included in the causal model, ABx, CS, PPIs, H2Bs, statins and NSAIDs were identified in this cohort. ABx were prescribed in 36% of patients within 28 days of the start of ICIs (Table 1). The most commonly prescribed ABx were β-lactams (**Figure S1**). CS were prescribed in 40% patients within 28 days of the start of ICIs. The most commonly prescribed CS were dexamethasone and prednisone (**Figure S2**). PPIs were prescribed in 37% of patients. Some patients received a single medication and no others during the study period, however, more frequently patients received several medications, e.g. CS with PPI and ABx, consistent with prophylaxis for developing an ulcer or pneumonia (**Figure S3**). The analysis strategy first tested for an association of a medication with OS without controlling for confounding effects of other medications and then further explored those medications with strong associations.

Across all cancer types, patients who were prescribed ABx within 28 days of the start of ICIs showed decreased OS (**Figure 2A**). This was also true of patients prescribed CS (**Figure 2B**), but not of patients prescribed other medications (**Figure 2C**). ABx showed a strong negative correlation with OS in RCC, NSCLC, melanoma, and bladder cancer. CS showed a strong negative correlation with OS in NSCLC, melanoma and other cancers. While other medications were not significantly associated with OS across all cancers, several showed significant associations with specific cancers. For example, H2Bs and NSAIDs associated with decreased OS in sarcoma and NSCLC, respectively. On the other hand, PPIs and Statins positively associated with OS in sarcoma. However, we observed the strongest associations for ABx and CS, and therefore followed these medications in further analyses.

**Figure 2.**
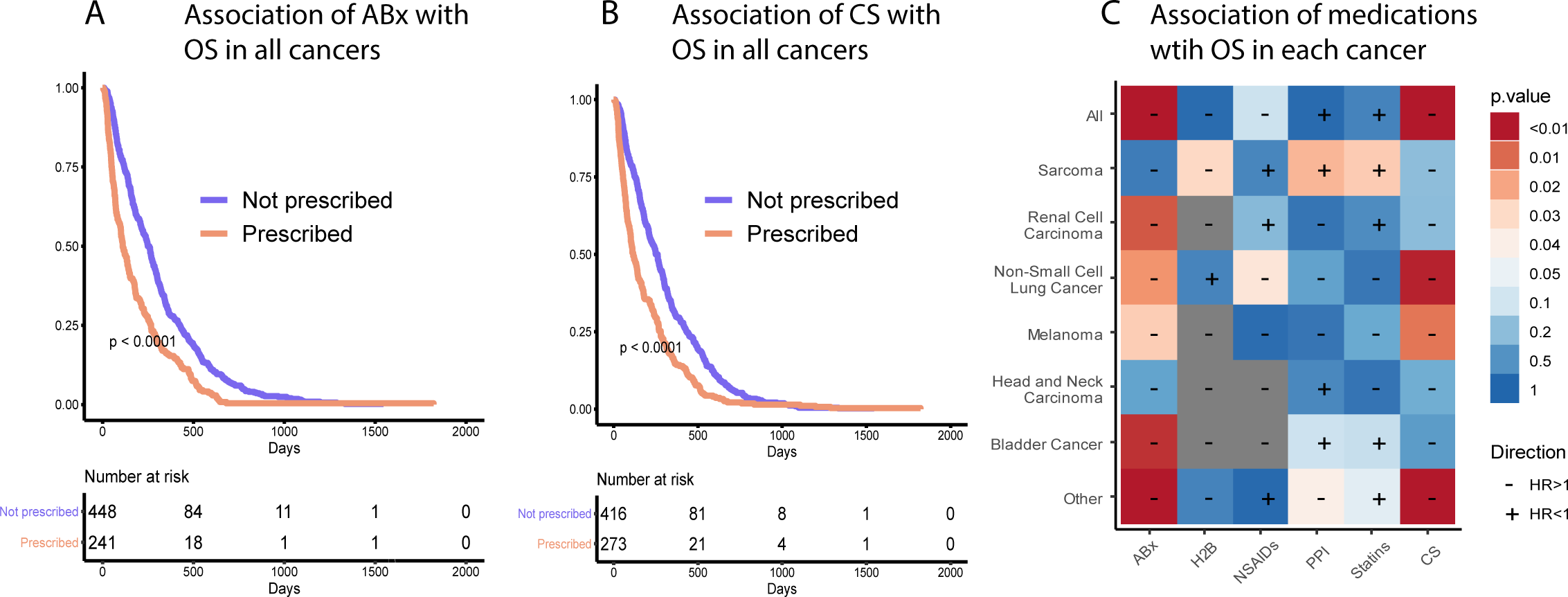
The effect of medications at the start of ICI treatment across all cancers for A) Antibiotics, B) Corticosteroids, and C) other medications. The cell color indicates the p-value of the Kaplan-Meier curve and the “+” or “-” the direction of the HR, in reference to its association with OS (i.e. a “-” indicates an association with decreased OS, therefore a HR > 1).

### Timing of medication use

Next we focused on the timing of ABx and CS prescriptions and their associations with OS in each cancer, using a 30-day sliding window (see Methods). ABx showed a greater HR than CS over nearly the entire period, and both were negatively associated with OS (**Figure 3A**). ABx treatment showed the highest HR more than 100 days before the start of ICIs, a second peak near day 50, and a third, lesser peak around day 0. CS showed a single, strong peak at day 0. Similar effects have been observed previously, but with little consistency in the time window tested (**Table 2**) (2,8,26–31). However, the majority of studies focused on a window at or near the start of ICI treatment. We therefore focused the timing analyses around ICI day 0 to capture the largest HR for both ABx and CS and to best compare the results to previous findings, and then examined the effects across cancers and drug subclasses.

**Table 2.**
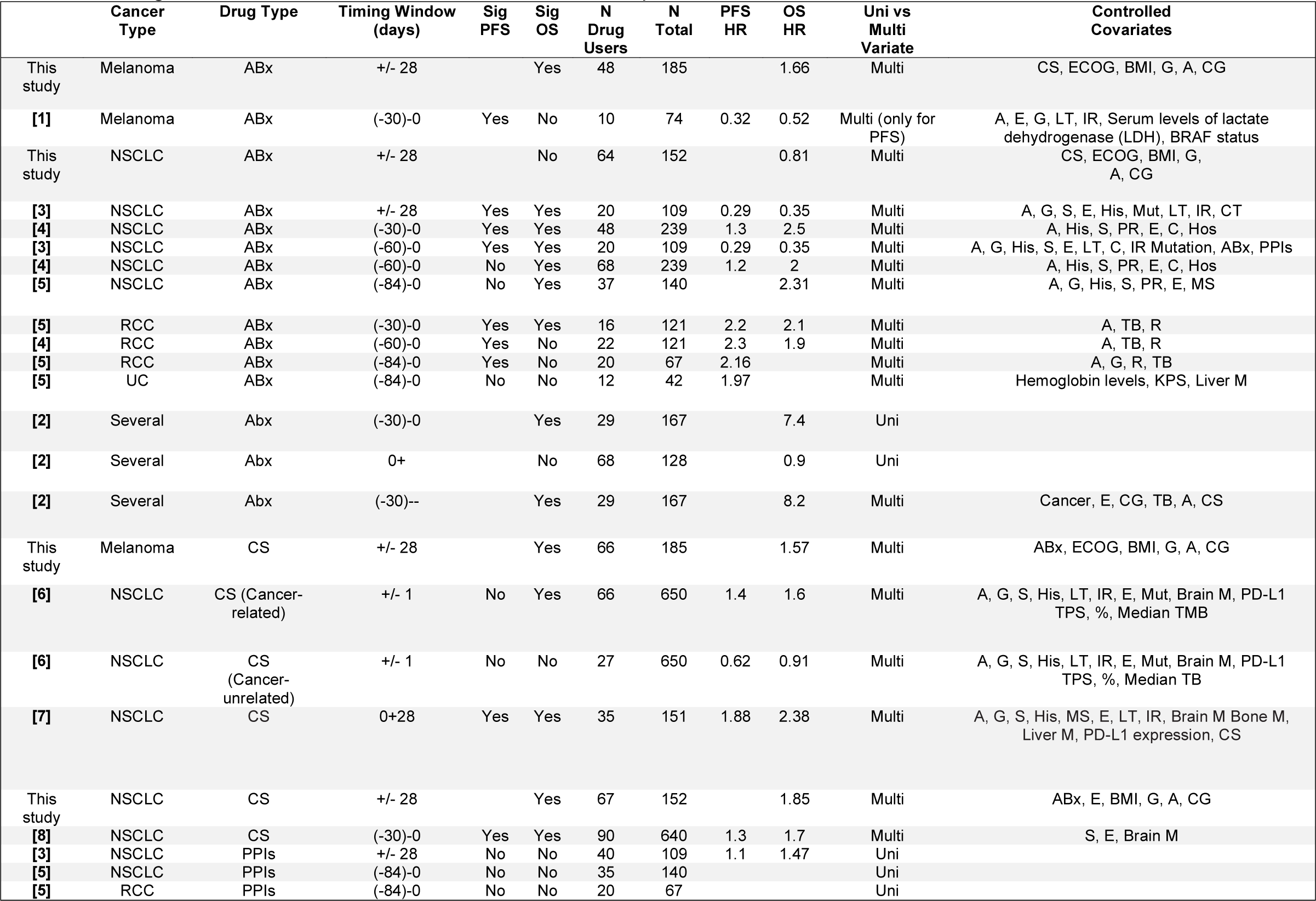
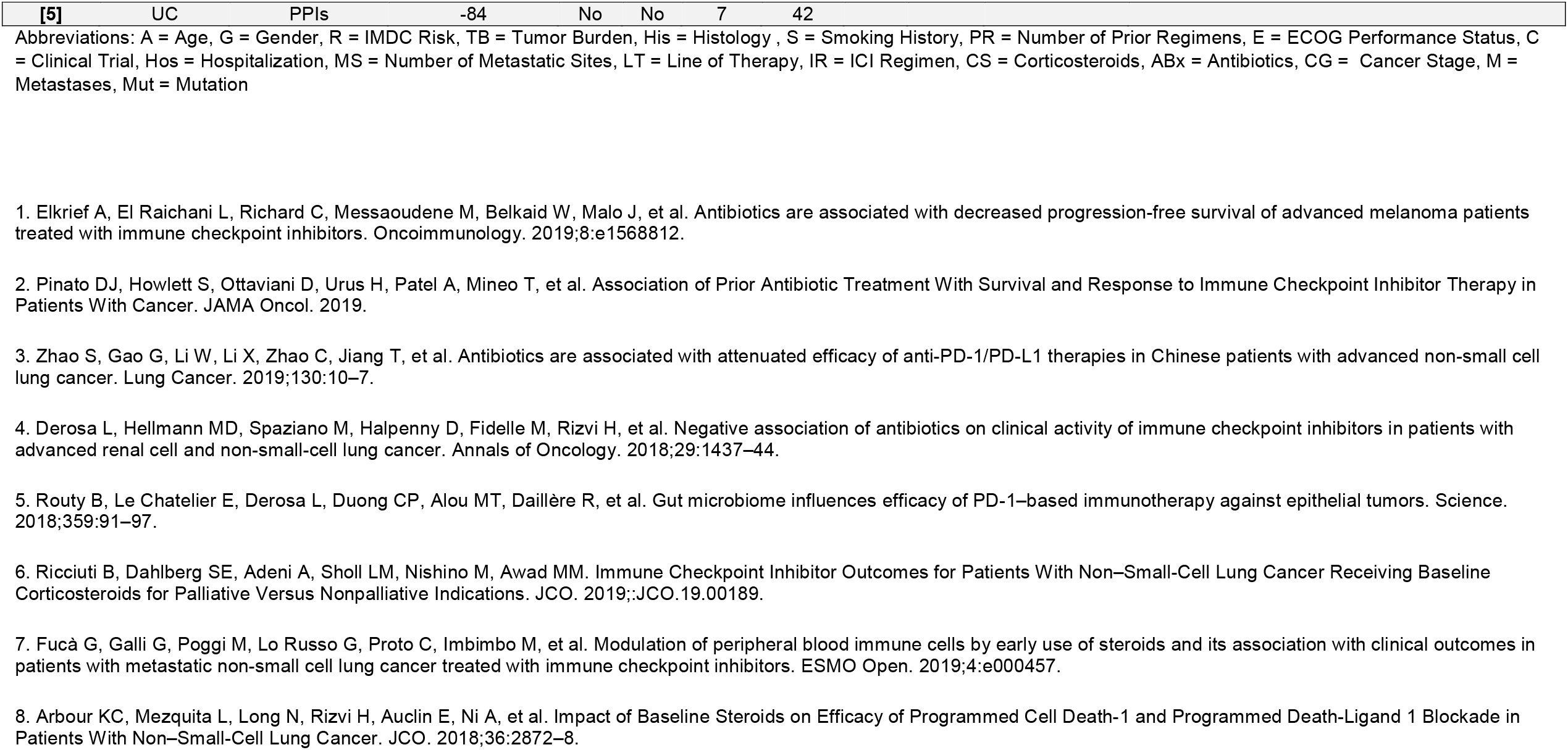
Timing of associations between medications and ICI response.

|  | Cancer Type | Drug Type | Timing Window (days) | Sig PFS | Sig OS | N Drug Users | N Total | PFS HR | OS HR | Uni vs Multi Variate | Controlled Covariates |
| --- | --- | --- | --- | --- | --- | --- | --- | --- | --- | --- | --- |
| This study | Melanoma | ABx | +/- 28 |  | Yes | 48 | 185 |  | 1.66 | Multi | CS, ECOG, BMI, G, A, CG |
| [1] | Melanoma | ABx | (-30)-0 | Yes | No | 10 | 74 | 0.32 | 0.52 | Multi (only for PFS) | A, E, G, LT, IR, Serum levels of lactate dehydrogenase (LDH), BRAF status |
| This study | NSCLC | ABx | +/- 28 |  | No | 64 | 152 |  | 0.81 | Multi | CS, ECOG, BMI, G, A, CG |
| [3] | NSCLC | ABx | +/- 28 | Yes | Yes | 20 | 109 | 0.29 | 0.35 | Multi | A, G, S, E, His, Mut, LT, IR, CT |
| [4] | NSCLC | ABx | (-30)-0 | Yes | Yes | 48 | 239 | 1.3 | 2.5 | Multi | A, His, S, PR, E, C, Hos |
| [3] | NSCLC | ABx | (-60)-0 | Yes | Yes | 20 | 109 | 0.29 | 0.35 | Multi | A, G, His, S, E, LT, C, IR Mutation, ABx, PPIs |
| [4] | NSCLC | ABx | (-60)-0 | No | Yes | 68 | 239 | 1.2 | 2 | Multi | A, His, S, PR, E, C, Hos |
| [5] | NSCLC | ABx | (-84)-0 | No | Yes | 37 | 140 |  | 2.31 | Multi | A, G, His, S, PR, E, MS |
| [5] | RCC | ABx | (-30)-0 | Yes | Yes | 16 | 121 | 2.2 | 2.1 | Multi | A, TB, R |
| [4] | RCC | ABx | (-60)-0 | Yes | No | 22 | 121 | 2.3 | 1.9 | Multi | A, TB, R |
| [5] | RCC | ABx | (-84)-0 | Yes | No | 20 | 67 | 2.16 |  | Multi | A, G, R, TB |
| [5] | UC | ABx | (-84)-0 | No | No | 12 | 42 | 1.97 |  | Multi | Hemoglobin levels, KPS, Liver M |
| [2] | Several | Abx | (-30)-0 |  | Yes | 29 | 167 |  | 7.4 | Uni |  |
| [2] | Several | Abx | 0+ |  | No | 68 | 128 |  | 0.9 | Uni |  |
| [2] | Several | Abx | (-30)-- |  | Yes | 29 | 167 |  | 8.2 | Multi | Cancer, E, CG, TB, A, CS |
| This study | Melanoma | CS | +/- 28 |  | Yes | 66 | 185 |  | 1.57 | Multi | ABx, ECOG, BMI, G, A, CG |
| [6] | NSCLC | CS (Cancer-related) | +/- 1 | No | Yes | 66 | 650 | 1.4 | 1.6 | Multi | A, G, S, His, LT, IR, E, Mut, Brain M, PD-L1 TPS, %, Median TMB |
| [6] | NSCLC | CS (Cancer-unrelated) | +/- 1 | No | No | 27 | 650 | 0.62 | 0.91 | Multi | A, G, S, His, LT, IR, E, Mut, Brain M, PD-L1 TPS, %, Median TB |
| [7] | NSCLC | CS | 0+28 | Yes | Yes | 35 | 151 | 1.88 | 2.38 | Multi | A, G, S, His, MS, E, LT, IR, Brain M Bone M, Liver M, PD-L1 expression, CS |
| This study | NSCLC | CS | +/- 28 |  | Yes | 67 | 152 |  | 1.85 | Multi | ABx, E, BMI, G, A, CG |
| [8] | NSCLC | CS | (-30)-0 | Yes | Yes | 90 | 640 | 1.3 | 1.7 | Multi | S, E, Brain M |
| [3] | NSCLC | PPIs | +/- 28 | No | No | 40 | 109 | 1.1 | 1.47 | Uni |  |
| [5] | NSCLC | PPIs | (-84)-0 | No | No | 35 | 140 |  |  | Uni |  |
| [5] | RCC | PPIs | (-84)-0 | No | No | 20 | 67 |  |  | Uni |  |

|  |  |  |  |  |  |  |  |
| --- | --- | --- | --- | --- | --- | --- | --- |
| **[5]** | UC | PPIs | -84 | No | No | 7 | 42 |

Abbreviations: A = Age, G = Gender, R = IMDC Risk, TB = Tumor Burden, His = Histology, S = Smoking History, PR = Number of Prior Regimens, E = ECOG Performance Status, C = Clinical Trial, Hos = Hospitalization, MS = Number of Metastatic Sites, LT = Line of Therapy, IR = ICI Regimen, CS = Corticosteroids, ABx = Antibiotics, CG = Cancer Stage, M = Metastases, Mut = Mutation

**Figure 3.**
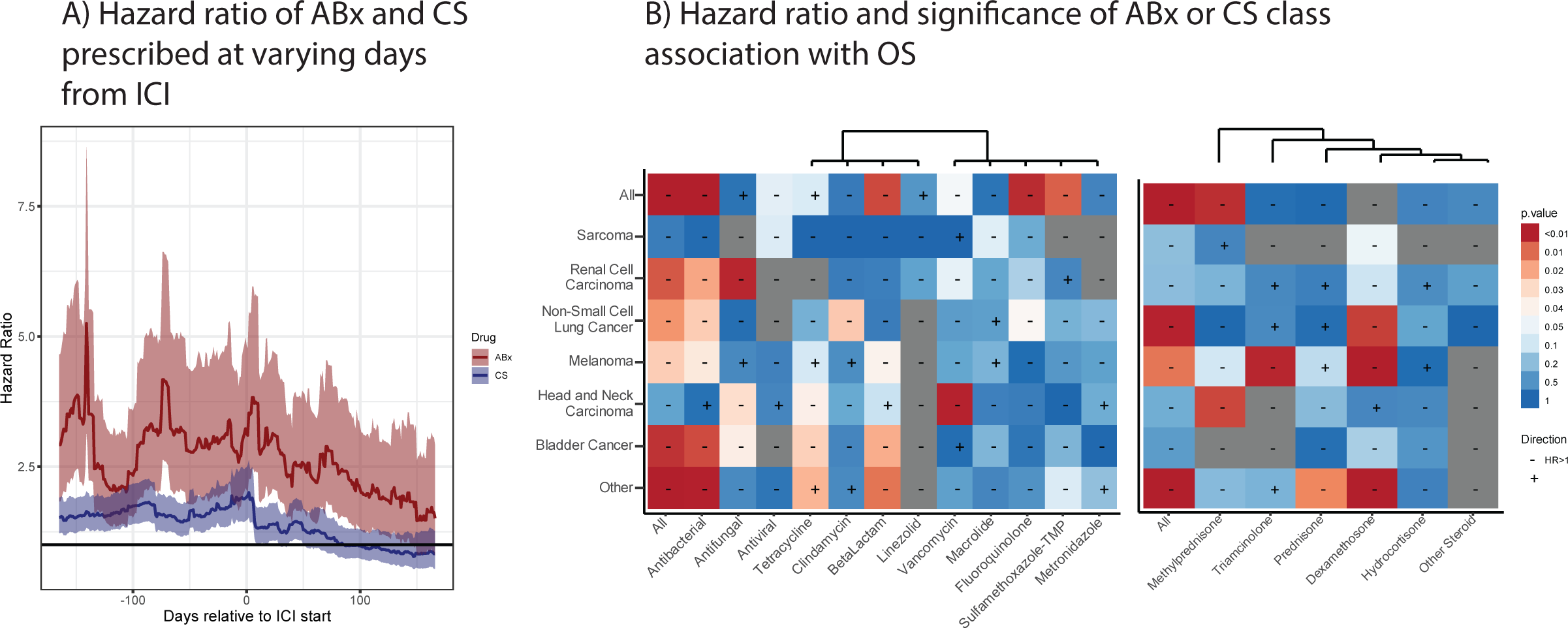
Associations of ABx and CS over time and by drug class. A) Hazard ratios with 95% confidence intervals of a Cox Proportional-Hazards model comparing individuals treated with ABx or CS during a 30-day sliding window compared to indivduals who did not receive ABx or CS, respectively. The significance and direction of associations of Cox Proportional Hazards models by (B) ABx or (C) CS class and cancer, using a window 28 days around ICI treatment start.

### Antibiotics and Corticosteroids classes

The effect of ABx on treatment response in different cancer types was not consistently associated with ABx class (**Figure 3B**). For example, β-lactams showed the highest HR in melanoma, but vancomycin (oral) had the highest HR in head and neck squamous cell carcinoma (HNSC). In addition, the overall effect of ABx was sometimes associated with a single ABx subclass and sometimes distributed over many. Additionally, NSCLC strongly associated with fluoroquinolone ABx, and this effect was stronger than the effect observed for all ABx combined. By contrast, the combined effect of all ABx in melanoma was much stronger than any individual ABx class. In fact, tetracycline was positively associated with OS in melanoma patients, despite the overall effect of ABx being negatively associated. On the other hand, the effects of CS classes on different cancers was more consistent (**Figure 3C**), though these comparisons were often limited by the sample size. Particularly with small sample sizes, confounding effects of patients receiving multiple drugs, e.g. ABx and CS, may dominate associations with OS. We therefore used combined models of ABx and CS to examine the effects of each.

### Combined modeling of ABx and CS, controlling for covariates

Models containing both ABx and CS showed that both are significantly associated with OS. A Kaplan-Meier curve stratifying patients by ABx, CS, or both, showed nearly identical intermediate effects of either ABx or CS, and an additive combined effect (**Figure 4A**). We next sought to control for confounding covariates using a Cox Proportional Hazards model. Including CCI, PS, BMI, sex, stage, and age in the model confirmed that ABx and CS remained highly significant, as were PS, BMI and age (**Figure 4B**). This suggests that ABx and CS are affecting OS in the context of ICI therapy by a mechanism other than that which is captured by PS, BMI or age, and is consistent with the microbiome parent to T-cell inflammation and child of ABx (**Figure 1**).

**Figure 4.**
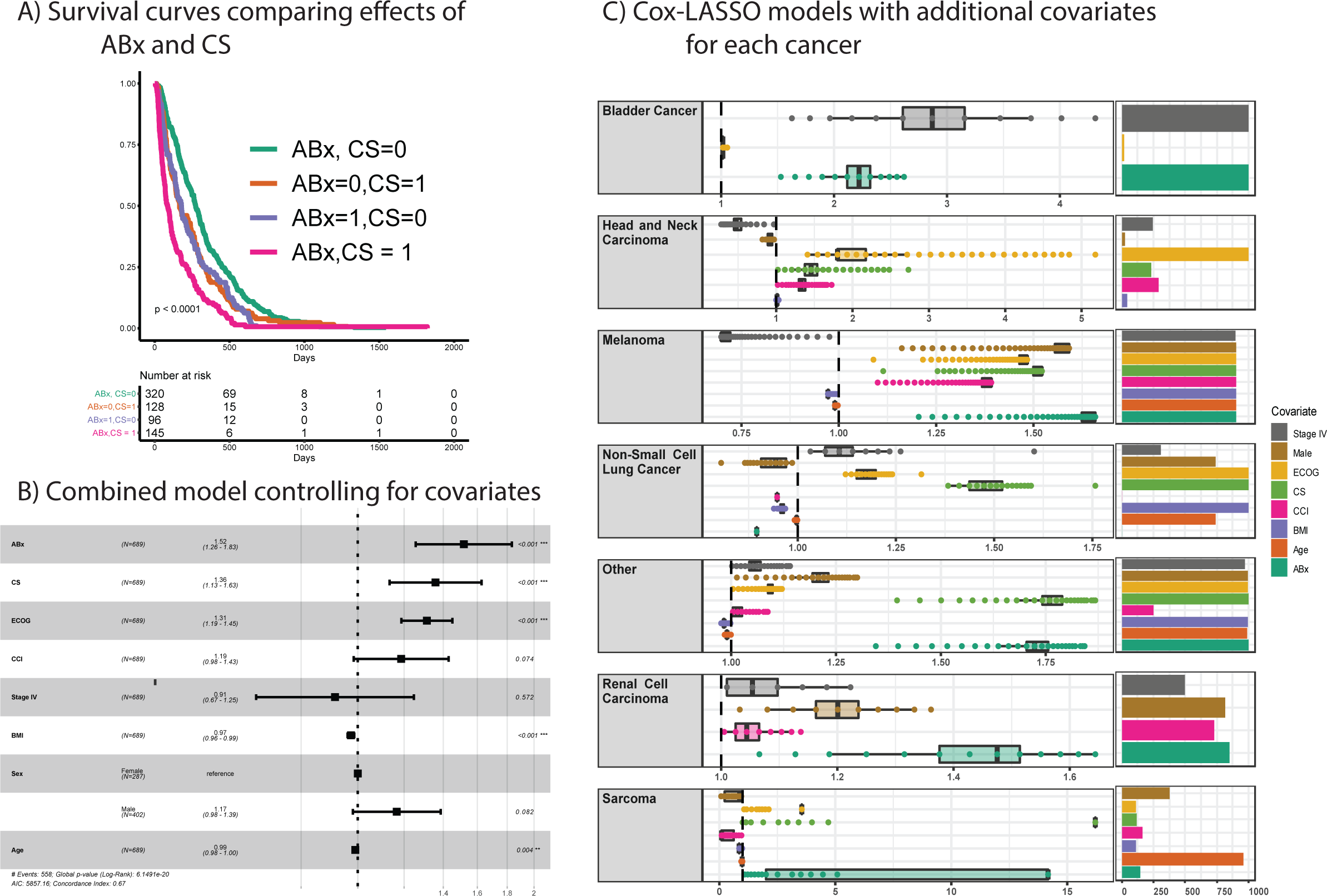
Combined models for ABx and CS and controlling for covariates. (A) Kaplan-Meier curves for ABx and CS in combination. B) Cox Proportional Hazards model incorporating both ABx and CS as well as several covariates. C) Cox-LASSO models for each cancer showing the hazard ratios estimated for covariates and the number of times the covariate was included in the model. The regularization parameter was selection by 10-fold cross validation, and then the robustness was assessed by 1000 bootstrap replicates using different random samples of the data.

In order to estimate the effects of ABx and CS within each cancer, we applied a method that (1) allowed different covariates to be included in each cancer, commensurate with the different clinical features of each cancer, and (2) removed uninformative variables, increasing the power for those cancers with smaller numbers of patients in this dataset. In addition, we repeated the analysis with different random samplings of the data in order to estimate the robustness of the variable selection. We found ABx to consistently and significantly associate with OS in bladder cancer, melanoma and RCC, but not in HNSC, NSCLC, or sarcoma. The HR was above 1 in each of the cancers where ABx was a consistently-selected covariate. Melanoma was notable in that all variables were consistently selected, with ABx showing the highest HRs (**Figure 4C**).

### The relationship between ABx, OS, and the microbiome

The bacterial taxa that showed the strongest enrichment in responders or non-responders to ICIs were selected from the literature and combined into a phylogenetic tree (**Figure 5**) (1,2,32). The taxa spanned several phyla and few ranks were consistently enriched in either responders or non-responders. For example, Firmicutes was found to be enriched in responders (1), but within the phylum are several taxa found to be enriched in non-responders (2,32). An exception to this was Bacteroidetes, which was found to be enriched in non-responders and each of the four species in the phylum were also enriched in non-responders (1,2). We performed a literature review of ABx susceptibilities for each of these taxa to estimate whether the size of the HR of the ABx would relate to the taxa for which it is active. For example, an ABx that target only bacteria enriched in non-responders may be beneficial because it may shift the community toward those taxa enriched in responders. On the other hand, if the overall diversity of the microbiome is important, broad-spectrum ABx may have higher HRs than narrow-spectrum.

**Figure 5.**
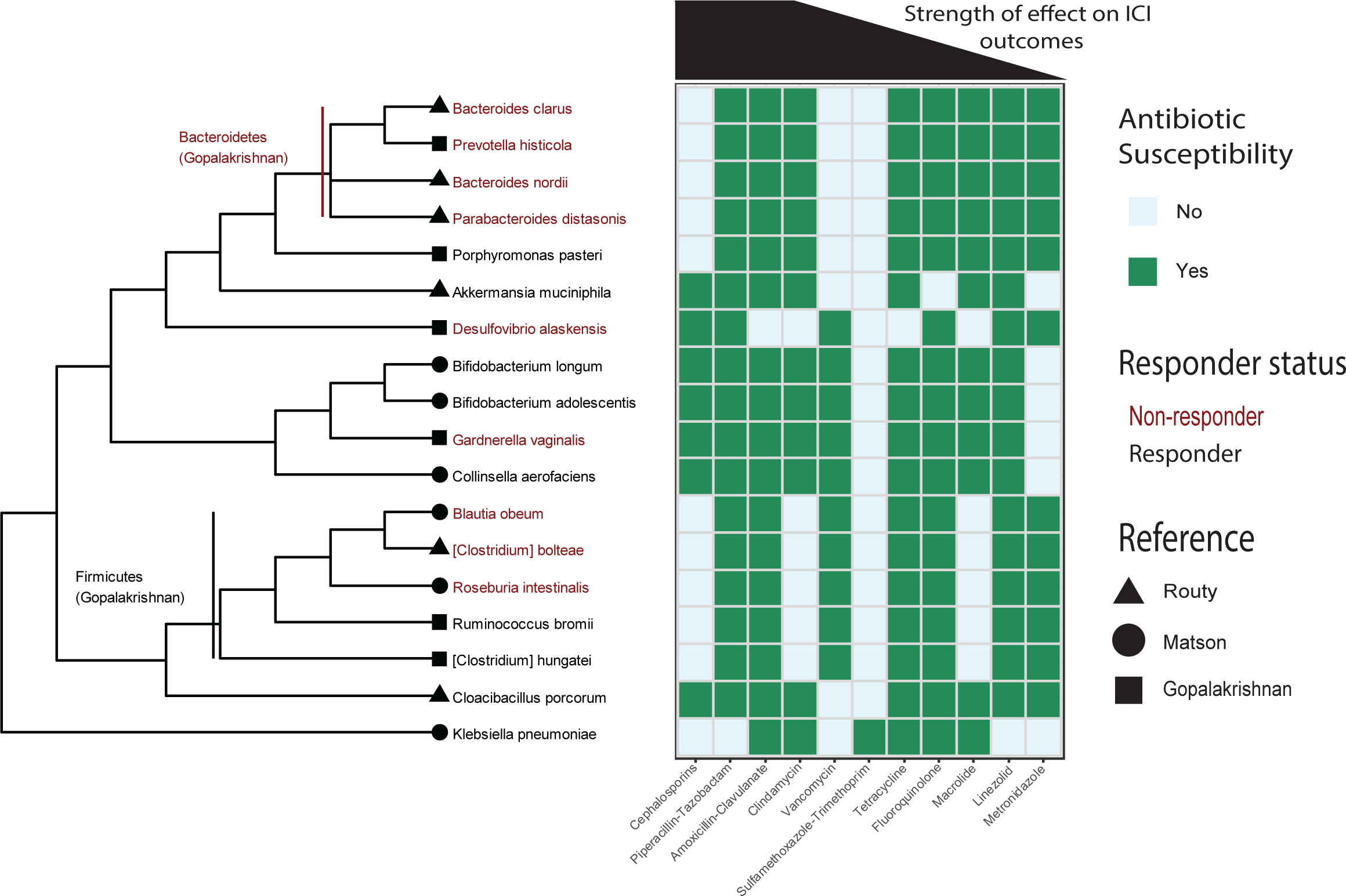
Relating the ABx effect to microbes enriched in responders to ICIs. A dendrogram of the microbes recently shown to be most enriched in responders (black) or non-responders (red), are related to known ABx susceptibilities (references for each cell in **Table S1**). The ABx are ordered by hazard ratio across all cancers (i.e. β-lactams showed the largest hazard ratio and linezolid the smallest).

The ABx class with the largest HR across all cancers was the β-lactams. Within this group category are the cephalosporins, which have a relatively narrow spectrum of activity and a unique pattern relative to other ABx classes. The cephalosporins are ineffective against the Bacteroidetes, found to be enriched in non-responders, but so were ABx such as vancomycin and sulfamethoxazole-trimethoprim (SXT). However, unlike vancomycin and SXT, cephalosporins effectively target *A. muciniphila*, which was shown to causally modify response to ICIs. Cephalosporins are also ineffective against several Firmicutes, similar to clindamycin, macrolides and metronidazole (**Figure 5**).

## DISCUSSION

The effects of medications or other variables are difficult to parse in a dynamic setting such as during treatment for cancer. We used a variety of methods to show that ABx and CS are significantly associated with decreased OS in several cancer types and that these results are consistent with mediation via the gut microbiome.

The association of CS with ICI response and OS remains controversial. Our observed association is consistent with other observations of decreased OS in NSCLC (8). However, Riccuiti et al showed no effect of CS on OS in NSCLC when given on the same day as ICI start, when the CS was prescribed for reasons other than “cancer-related palliative indications” (33). Our records lack some variables needed to replicate those results, however our results are consistent with aspects those findings. For example, dexamethasone treatment showed a strong negative association with OS across several cancer types, consistent with its use for brain metastases and anorexia, which are all indicators of poor clinical outcome. On the other hand, several of our analyses demonstrated associations between CS and OS that may not be due to selecting a sub-cohort with a poor prognosis. Our first causal strategy, the time analysis, showed similar results when restricting CS medications to a single day, but a larger effect when a wider time window was used (**Table 2**). Our second causal strategy, controlling for covariates, cannot be directly compared because our dataset did not include central nervous system metastases. However, when we control for metastatic stage and PS, the CS association remains. Our third causal strategy, comparisons between cancers, shows that the CS association with OS is observed in cancers for which brain metastases are not common, such as RCC, and for specific CS not typically used for brain metastases, such as methylprednisolone in HNSC. This suggests that understanding the association between CS and the response to ICIs may require more granular assessment of CS types (i.e. rather than collapsing to 10 mg prednisone equivalent) and cancers.

We applied the same logical framework to ABx treatment to demonstrate an effect on OS. Unlike CS, the majority of studies have found an association between ABx use and ICI response, independent of the time window (**Table 2**). Our longitudinal analysis showed a global maximum HR well before the start of ICIs, consistent with the ABx effects persisting for long periods. Given this result, it is unlikely that acute illnesses drive the association between ABx and OS. However, a recent prospective study found that ABx given currently with ICI treatment did not significantly affect OS for a group of patients with lung, skin, or several other cancers (26) (**Table 2**). We observe lower HRs for the effect of ABx after ICI start, however it remains significant until approximately 120 days post ICI start. We note that within cancers the effect of ABx is highly variable (**Figure 4C**); the difference may be due to the composition of the cohorts (e.g. more patients with bladder cancer, where ABx has a strong effect, and fewer with NSCLC, where the effect is less).

When controlling for illness-related covariates that report on the overall health status of the individual (e.g. CCI, PS) the effect of ABx remained significant. Third, the associations of ABx and OS were observed across cancer types (e.g. patients with bladder cancer versus melanoma). A larger fraction of bladder cancer patients were treated with ABx than any other cancer (56%), consistent with their use for urinary tract or as prophylaxis for invasive urologic procedures. On the other hand, melanoma patients treated with ABx were the smallest fraction of any cancer (25%), consistent with this population being less likely to undergo procedures in which prophylactic ABx are used. It is reasonable to suspect that melanoma patients treated with ABx are therefore more compromised than those not treated with ABx. However, an effect of ABx remains, even for bladder cancer. Although it remains probable that the cohorts who receive ABx are different from those who did not in ways that have not been controlled for in analyses, these three analyses add confidence to the association of ABx with OS in the context of ICIs.

We next related the strength of the association of ABx classes with OS and the microbes that those ABx classes affect. The β-lactam ABx were shown to have the strongest association with OS across cancer types. The literature review of antibiotic susceptibilities showed that this diverse class is effective against the Gram-positive phylum Firmicutes. The literature review of the bacterial taxa associated with response to ICIs, showed that the Firmicutes are enriched in responders to ICIs. Moreover, β-lactams are not consistently effective against members of the phylum Bacteroidetes, which was found to be enriched in non-responders. This suggests that the β-lactams may show the strongest signal across all cancers in our dataset because they disrupt the microbiome in such a way that they reduce response to ICIs by depleting the Firmicutes more so than the Bacteroidetes.

The association between ABx prescriptions and OS that we observe is consistent with direct measurements of the microbiome and response to ICIs (1–3). However, there is no consensus for which taxa are enriched in the responders to ICIs (**Figure 5**). For example, there is causal evidence for *Akkermansia muciniphila* increasing response to ICIs, however, it was not among the most enriched in the other datasets (1,2,32). Nonetheless, agreement can be observed at broader taxonomic characterizations. Non-responders are enriched for taxa in the phylum Bacteroidetes and this is consistent with the strength of association we see for different classes of ABx (1,2). Narrow spectrum β-lactams (e.g. cephalosporins), which show the strongest association with OS, are effective against Firmicutes (enriched in responders) but less so against Bacteroidetes (enriched in non-responders).

The results presented here contrast with several assumptions gathered from the literature and described by the causal model (**Figure 1**). First, we found that ABx and CS are the only medications significantly associated with OS, despite the inclusion of several medications associated with changes to the microbiome (**Figure 2**). This may be due to the types of changes incurred (e.g. PPIs may not significantly change the abundances of those taxa linked to ICI response) or the strength of the effect amid the noise in the data. However, the other two hypotheses were borne out by the analyses.

The CS and ABx medications showed an additive effect on OS, consistent with a collider interaction in the model (**Figure 4A**). Also, there was an effect of ABx after controlling for many covariates, consistent with its direct effect on the microbiome and the microbiome playing a role in ICIs (**Figure 4B**). This result was consistent with the relationship between the strength of the ABx signal and the bacterial taxa susceptible to that ABx (**Figure 5**).

### Limitations

A key challenge in this and other retrospective analyses is inferring causal relationships in non-randomized cohorts. For example, patients who receive medications such as antibiotics may be quite different from those who do not. However, it is difficult to imagine an ethical trial that could randomize treatment with ABx in this setting. Therefore, retrospective analyses may be the best option until direct measurements of the microbiome are widely available. We used a variety of methods to show that ABx and CS are significantly associated with decreased OS across a variety of cancers and that these results are consistent with a role for the gut microbiome.

Our study remains limited by being unable to account for important factors known to affect OS in the context of ICI treatment. For example, the complete ABx history of patients -- much longer than the windows reported here -- are very likely of consequence. Several groups have studied the recovery of microbiome diversity following ABx exposure and results show reasonable recovery 90 days later (34,35). However, multiple courses of ABx prevented such a recovery; i.e. diversity returned to baseline after one treatment with ABx, but not after a second ABx treatment within 60 days (36). It is therefore possible that individuals who show extreme effects of ABx treatment are beyond the time scale of this study. Without baseline microbiome diversity measures we are unable to capture such information. Similarly, estimating the effects of ABx on communities from data on microbes in isolation is, at best, approximate. A better understanding of how ABx affect complex communities is needed. Other limitations include our small sample size relative to the heterogeneity in the data. Future directions should capture variables such as the presence of brain metastases, tumor biomarkers such as tumor mutational burden and PD-1/PDL-1 status, and outcome variables like ICI response or the number of tumor-infiltrating lymphocytes.

## CONCLUSIONS

ABx and CS, but not other medications known to affect the microbiome, are associated with reduced OS when administered near the start of ICI treatment. Our results show this finding several cancer types, and for several subclasses of these drugs. These results are consistent with a role of the microbiome in response to ICIs and identify clinical settings where the microbiome is likely to play the largest role, namely NSCLC, melanoma, RCC, HNSC, and bladder cancer. A clear understanding of which microbes are important for ICI responses and in what cancers will require the collection of microbiome samples across a wide variety of clinical settings. However, some information can be gathered by indirect means, which identifies the settings where the microbiome is likely to have the greatest effects. Medications that affect the microbiome given concomitantly with ICIs provide evidence for where microbes play a role. Further work is needed to identify which microbes are important and identify solutions to mitigate these effects and perhaps promote greater response to ICIs.

## LIST OF ABBREVIATIONS

ICIs: Immune Checkpoint Inhibitors
OS: Overall Survival
ABx: Antibiotics
CS: Corticosteroids
CCI: Charlson Comorbidity Index
PS: Eastern Cooperative Oncology Group Performance Status
BMI: Body Mass Index
NSCLC: Non-Small Cell Lung Cancer
RCC: Renal Cell Carcinoma
HNSC: Head and Neck Squamous Cell Carcinoma

## DECLARATIONS

### Ethic Approval and Consent to Participate

Data from this work were collected under an IRB-approved retrospective study (OSU-2016C0070, OSU-2017C0063).

### Consent for Publication

Not applicable.

### Author’s Contributions

MH, GT, SP, JTB, CFV, KLK, SH, JP, DQ, GAO, MG, CP and DHO collected the data. RH, MM, JB, LW, LL, DPC, DHO and DS analyzed the data and generated the figures and tables. DS, RH and DHO wrote the manuscript. All authors read and approved the final manuscript.

## Acknowledgements

This work was performed on the Ohio Supercomputer Center servers (Owens, Pitzer; Project PA1490 awarded to Daniel Spakowicz). The authors would like to thank Stephanie Fortier (Division of Medical Oncology, The Ohio State University; ORCID 0000-0002-2380-2378) for critically reviewing and editing the manuscript.

## SUPPLEMENTARY FIGURES AND TABLES

**Figure S1. Causal diagram with references**.

**Figure S2. Number of medications prescribed across all cancers and the frequency of multiple medications**.

**Figure S3. Number of antibiotics, separated by class, prescribed across all cancers and the frequency of multiple antibiotics**.

**Figure S4. Number of corticosteroids, separated by class, prescribed across all cancers and the frequency of multiple antibiotics**.

**Table S1. References for the antibiotic susceptibilities of bacterial taxa found to be enriched in responders or non-responders to ICI therapy**

